# Non-inferiority of a red-blood-cell–only transfusion strategy compared with balanced resuscitation in adults with massive gastrointestinal haemorrhage: a propensity-score–weighted cohort study

**DOI:** 10.64898/2026.05.25.26354037

**Authors:** Burak Bahar, Joseph D. Sweeney, Christian Nixon

## Abstract

**Background:** Balanced (1:1:1) transfusion of red blood cells (RBCs), plasma, and platelets is the standard of care in trauma-induced massive haemorrhage, where early coagulopathy is a defining feature. In gastrointestinal (GI) haemorrhage this physiology is non-prominent, and whether plasma and platelets provide benefit when ≥ 10 RBC units are required within 24 hours is unknown.

**Objective:** To test whether a red-blood-cell-only (RBC-only) transfusion strategy is non-inferior to a balanced (Balanced) strategy for in-hospital mortality in adults meeting massive-transfusion criteria for GI haemorrhage.

**Design:** Single-centre retrospective cohort of 559 adult massive-transfusion encounters (536 patients; 2021–2025) with a primary admitting diagnosis of upper, lower, or unspecified GI haemorrhage. Exposures were RBC-only versus Balanced (RBCs with any plasma and/or platelets). The primary outcome was in-hospital mortality, with a pre-specified 5-percentage-point (pp) non-inferiority margin on the absolute risk difference and a 3-pp sensitivity margin. Analysis used augmented inverse-probability-of-treatment weighting (AIPTW) with bootstrap inference (2,000 resamples by patient). Five pre-specified sensitivity analyses were performed.

**Results:** 505 encounters (90.3%) received RBC-only and 54 (9.7%) received Balanced transfusion. The AIPTW risk difference for in-hospital mortality (RBC-only − Balanced) was -19.8 pp (95% CI -68.1 − -2.2 pp). Non-inferiority was demonstrated at both the primary 5-pp and the more stringent 3-pp margins. Five pre-specified sensitivity analyses, (1) a propensity-score matched cohort, (2) a complete-case model incorporating INR, (3) a broader GI diagnosis set (n = 749), (4) a first encounter per patient restriction, and (5) E-value bound analysis were concordant with the primary estimate.

**Conclusion:** In this propensity-score–weighted cohort of adults with massive GI haemorrhage, an RBC-only transfusion strategy was non-inferior to a balanced strategy for in-hospital mortality at both 5-pp and 3-pp margins. The findings support individualized use of plasma and platelets in GI haemorrhage rather than reflexive application of the 1:1:1 trauma protocol; prospective confirmation is warranted.

**Significance of this study:** *What is already known on this topic:* Balanced 1:1:1 transfusion is the standard of care for trauma-induced massive haemorrhage, where early coagulopathy, hypothermia, and acidosis frequently coexist. Whether this model should be extrapolated to non-traumatic gastrointestinal haemorrhage, a population in which the trauma triad is generally absent, is uncertain. To date, randomized trials in acute GI bleeding tested restrictive RBC thresholds and antifibrinolytic therapy but have not directly compared red-cell-only with balanced component strategies in patients meeting massive-transfusion criteria.^1–3^

*What this study adds:* In a propensity-score–weighted cohort of 559 adults meeting massive-transfusion criteria for GI haemorrhage, an RBC-only strategy was non-inferior to a balanced strategy for in-hospital mortality at both the pre-specified 5-pp margin and a more stringent 3-pp sensitivity margin. The finding was consistent across all pre-specified sensitivity analyses, including 1:1 propensity-score matching, complete-case adjustment for admission INR, a broader GI-diagnosis cohort, a first encounter per patient restriction, and an E-value bound for unmeasured confounding.

*How this study might affect research, practice or policy:* These results support individualized use of plasma and platelets rather than universal and reflexive 1:1:1 transfusion in GI haemorrhage. The results support a randomized trial comparing goal-directed component therapy with a fixed-ratio resuscitation in non-traumatic massive GI bleeding.

## Introduction

The management of haemorrhage control has been transformed over the past century, evolving from crystalloid resuscitation, through whole-blood transfusion, to component-based protocols that intentionally limit crystalloid use.^4–6^ Traditional massive transfusion protocols (MTPs) administered red blood cells as the primary resuscitative component, with plasma and platelets added subsequently and reactively once overt dilutional coagulopathy had developed. Damage control resuscitation (DCR; also termed balanced or algorithm-driven resuscitation), arising from military trauma practice, reverses this sequence: plasma and platelets are given pre-emptively, in a fixed ratio with RBCs, to forestall the coagulopathy that would otherwise develop with large-volume resuscitation, alongside permissive hypotension and early haemorrhage control. DCR is the resuscitation strategy; an MTP is the operational delivery system that makes rapid, ratio-based component access feasible.^6–8^ In trauma the two are bundled and this bundle has been associated with improved survival at 72 hours in selected trauma patients.^9^ Massive transfusion (MT), commonly defined as ≥ 10 units of RBCs within 24 hours,^7, 10^ is required in a minority of patients admitted with gastrointestinal (GI) haemorrhage but accounts for disproportionate blood-product utilisation.^11^

In trauma, the PROPPR trial established that a 1:1:1 ratio of plasma, platelets, and RBCs accelerates haemostasis and reduces death from exsanguination at 24 hours compared with a 1:1:2 strategy.^9^ Balanced resuscitation has since become the default practice for institutional massive-transfusion protocols, including in non-trauma settings such as GI haemorrhage, however, the paucity of evidence and physiological rationale is considerably weaker and no mortality difference were found at different plasma to RBC transfusion ratios.^12, 13^ Furthermore, GI bleeding patients are a heterogeneous population ranging from young patients with peptic ulcer to elderly with cirrhosis and portal hypertension.

The coagulopathy that justifies early, empirical plasma and platelet support in trauma is a distinct clinical entity: it arises from tissue injury, shock, and hyperfibrinolysis and is present in a substantial fraction of severely injured patients on arrival.^4, 8, 14, 15^ GI haemorrhage, by contrast, rarely presents with the triad of coagulopathy, hypothermia, and acidosis, and the coagulation disturbance when present is more commonly attributable to pre-existing liver disease, anticoagulant therapy, or dilutional effects of large-volume RBC resuscitation rather than to acute consumption.^13, 16, 17^ It is therefore unclear whether empirically administering plasma and platelets confers benefit in the GI haemorrhage setting, or whether it simply exposes patients to allogeneic component products and their attendant transfusion-reaction, volume-overload, and infectious risks especially for patients with portal hypertension where aggressive plasma transfusion can increase portal venous pressure, potentially exacerbating active bleeding or causing re-bleeding and for patients who are prone to developing transfusion associated circulatory overload.^18–20^ This scepticism is reinforced by the HALT-IT trial, in which tranexamic acid, a common trauma intervention, conferred no mortality benefit and increased venous thromboembolism and seizures in acute GI bleeding.^3^ A nationwide study of over 5,000 patients with acute non-variceal GI bleeding showed a high ratio of Balanced transfusion reduced the need for re-endoscopy but was associated with increased mortality.^21^ These studies showed that trauma-validated haemostatic interventions do not automatically transfer to GI-bleed physiology.

A non-inferiority design directly addresses the policy-relevant question: can plasma and platelets be safely withheld in massive GI haemorrhage? We pre-specified an absolute risk-difference margin of 5 percentage points on in-hospital mortality which is close to the margins used in the previous transfusion-threshold trials and with a more stringent 3-pp margin for sensitivity.^1, 2, 22, 23^ We tested the hypothesis that an RBC-only strategy is non-inferior to a balanced strategy in adults meeting MT criteria for GI haemorrhage, using augmented inverse-probability-of-treatment weighting (AIPTW) to adjust for confounding by indication across an 10-covariate propensity model.^24–26^

## Methods

### Study design and data source

This was a retrospective single-centre cohort study, designed as a non-inferiority analysis of a binary transfusion exposure on in-hospital mortality. The analysis was conducted under umbrella IRB protocol 2339166-1 (“Design and implementation of a transfusion medicine data science platform”), effective 2025-10-01, and was pre-specified as a sub-analysis of that protocol; no separate IRB submission was required. The completed STROBE checklist is provided. The STROBE statement^27^ was used to guide observational reporting and the CONSORT-NI extension^28^ was used to inform reporting of the non-inferiority components. The analytic dataset comprised all adult inpatient encounters from 2021-01-01 through 2025-12-31 with structured admission, laboratory, transfusion, and outcome fields extracted from the institutional electronic health record.

### Cohort definition

Inclusion criteria were (1) age ≥ 18 years at admission, (2) inpatient status, (3) a primary admitting diagnosis of upper (UGIB), lower (LGIB), or unspecified GI haemorrhage, and (4) transfusion of ≥ 10 RBC units within 24 hours of admission. Repeat encounter contributions from the same patient were retained in the primary analysis and handled by cluster-robust inference. A sensitivity analysis restricted to each patient’s first eligible MT encounter is also reported (see sensitivity analyses).

### Exposure and outcomes

Patients were considered “RBC-only” if exposed to no plasma or platelet transfusions over the encounter or “Balanced” if either plasma and/or platelet transfusions occurred over the encounter.

Because the electronic extract reports plasma and platelet units as whole-encounter cumulative totals rather than 24-hour counts, the ratio-based exposure definitions used in trauma MT literature could not be computed on the acute window; the implications of this limitation are quantified in the limitations. The primary outcome was in-hospital mortality and secondary outcomes were ICU admission and length of stay (LOS).

### Non-inferiority margin

The pre-specified primary non-inferiority margin was an absolute risk difference of 5 percentage points (RBC-only − Balanced) on in-hospital mortality, evaluated as the upper one-sided 97.5% confidence bound below 5 pp. A 3-pp sensitivity margin was also pre-specified. The 5-pp primary margin is conservative against the 2–6 pp range conventional in transfusion-threshold and critical-care non-inferiority trials^1, 2, 22, 23^ and is a clinically appropriate threshold given the observed 1–15% absolute mortality range across the two exposure arms.

### Data cleaning

Admission creatinine values in the source extract showed evidence of non-serum sample contamination hence creatinine was excluded from the analyses.

### Statistical analysis

The primary estimator was the average treatment effect (ATE) on the risk difference for in-hospital mortality, obtained by augmented inverse-probability-of-treatment weighting (AIPTW), a doubly robust estimator that is consistent if either the propensity-score model or the outcome regression is correctly specified.^26, 29^ The propensity for Balanced versus RBC-only exposure was modelled with logistic regression on 10 pre-specified covariates, each with < 2% missingness in the cohort: age at admission, sex, first haemoglobin, 24-hour haemoglobin nadir, first platelet count, first blood urea nitrogen (BUN), variceal bleed, endoscopic intervention, RBC units within 24 hours, and admitting diagnosis subtype (UGIB/LGIB/Unspecified GI). Continuous covariates were entered as restricted cubic splines with 3 knots at the 10th, 50th, and 90th percentiles.

Weights were 1/ê for Balanced observations and 1/(1−ê) for RBC-only, where ê is the fitted propensity. To limit the influence of extreme weights, weights in the upper 1% were winsorised at the 99th percentile. Covariate balance pre- and post-weighting is reported as standardised mean differences (SMDs); the threshold |SMD| < 0.10 indicates acceptable balance.^30^ A 6-covariate outcome regression (logistic link on mortality; gamma-log link on LOS) using first haemoglobin, 24-hour haemoglobin nadir, age, first platelet, variceal bleed, and admitting diagnosis subtype provided the AIPTW augmentation.

Inference used a non-parametric cluster bootstrap with 2,000 resamples, resampling patients with replacement to respect within-patient correlation. 95% confidence intervals were percentile-based; non-inferiority was decided by the upper 97.5% confidence bound relative to the pre-specified 5-pp and 3-pp margins. A fixed seed was used throughout for reproducibility. All analyses were conducted in Python 3.10 (pandas 2.3, statsmodels 0.14, scipy 1.15) and independently replicated in R 4.5 (WeightIt 1.7, MatchIt 4.7, sandwich 3.1, boot 1.3); point and interval estimates were required to match between implementations within Monte Carlo noise.

### Sensitivity analyses

Five sensitivity analyses were pre-specified: (1) one-to-one propensity-score matching with caliper 0.2 × SD of the logit propensity, (2) complete-case AIPTW including first INR as an additional covariate, (3) broader GI admitting diagnosis inclusion via a regular expression applied to the free text admitting diagnosis field, (4) first eligible MT encounter per patient, and (5) E-value for the observed adjusted risk difference.^31^

### Patient and public involvement

This retrospective analysis of de-identified electronic-health-record data did not involve patients or the public in the design, conduct, reporting, or dissemination plans of the study.

## Results

### Cohort characteristics

Of 20,009 inpatient encounters in 2021–2025, 559 met pre-specified inclusion criteria, contributed by 536 unique patients. 505 encounters (90.3%) received an RBC-only strategy and 54 (9.7%) received Balanced transfusion. Baseline characteristics are reported in Table 1. Relative to the RBC-only arm, the Balanced arm was younger (median 68 vs 74 years, p<0.001), had a lower platelet count (140 vs 229 × 10⁹/L, p<0.001), a higher prevalence of variceal bleeding (35.2% vs 11.9%, p<0.001), received more RBC units within 24 hours (median 15 vs 12, p=0.004), and had substantially higher unadjusted in-hospital mortality (14.8% vs 1.2%, p<0.001) and ICU utilisation (27.8% vs 3.4%, p<0.001).

**Table 1.**
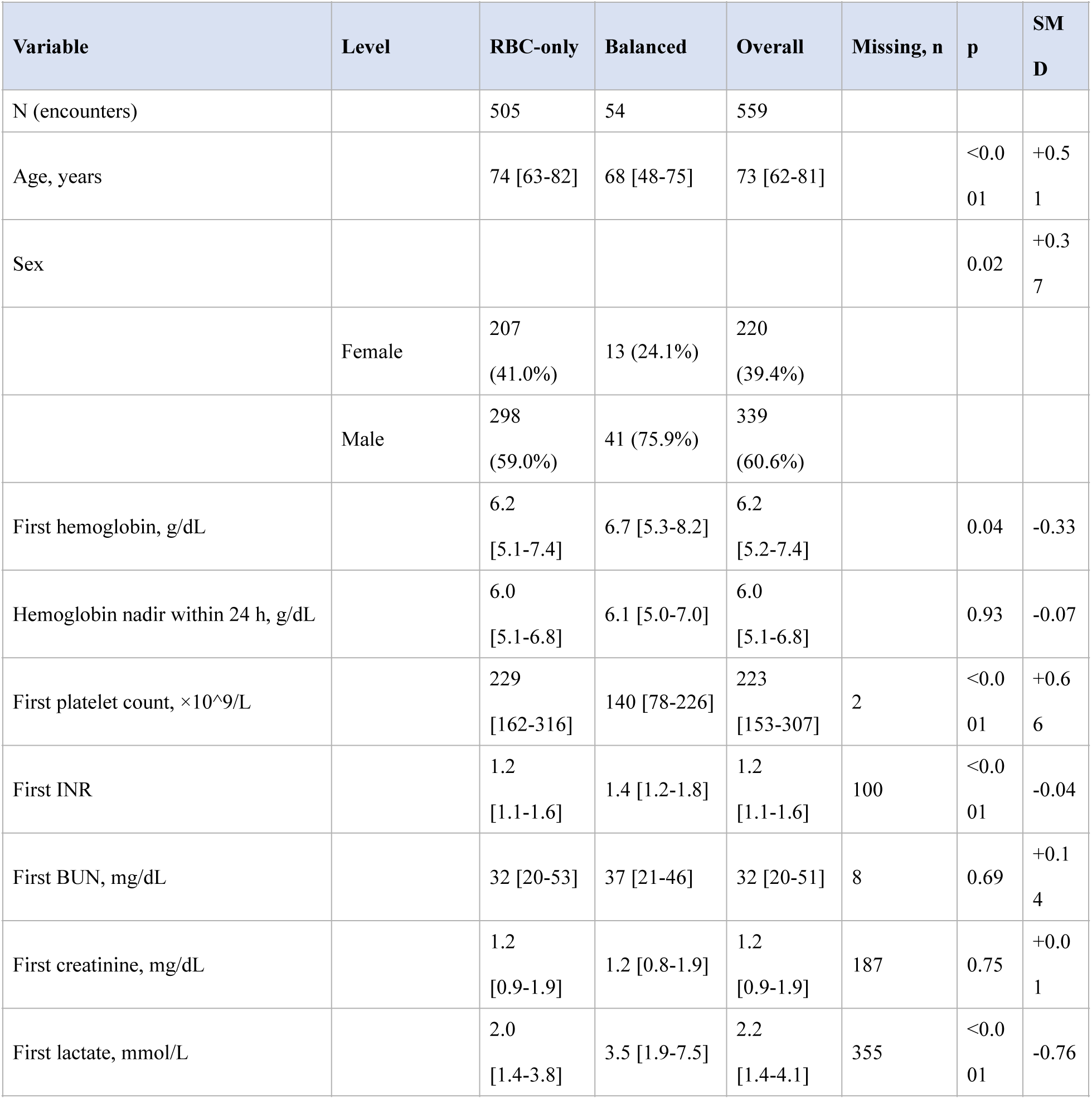
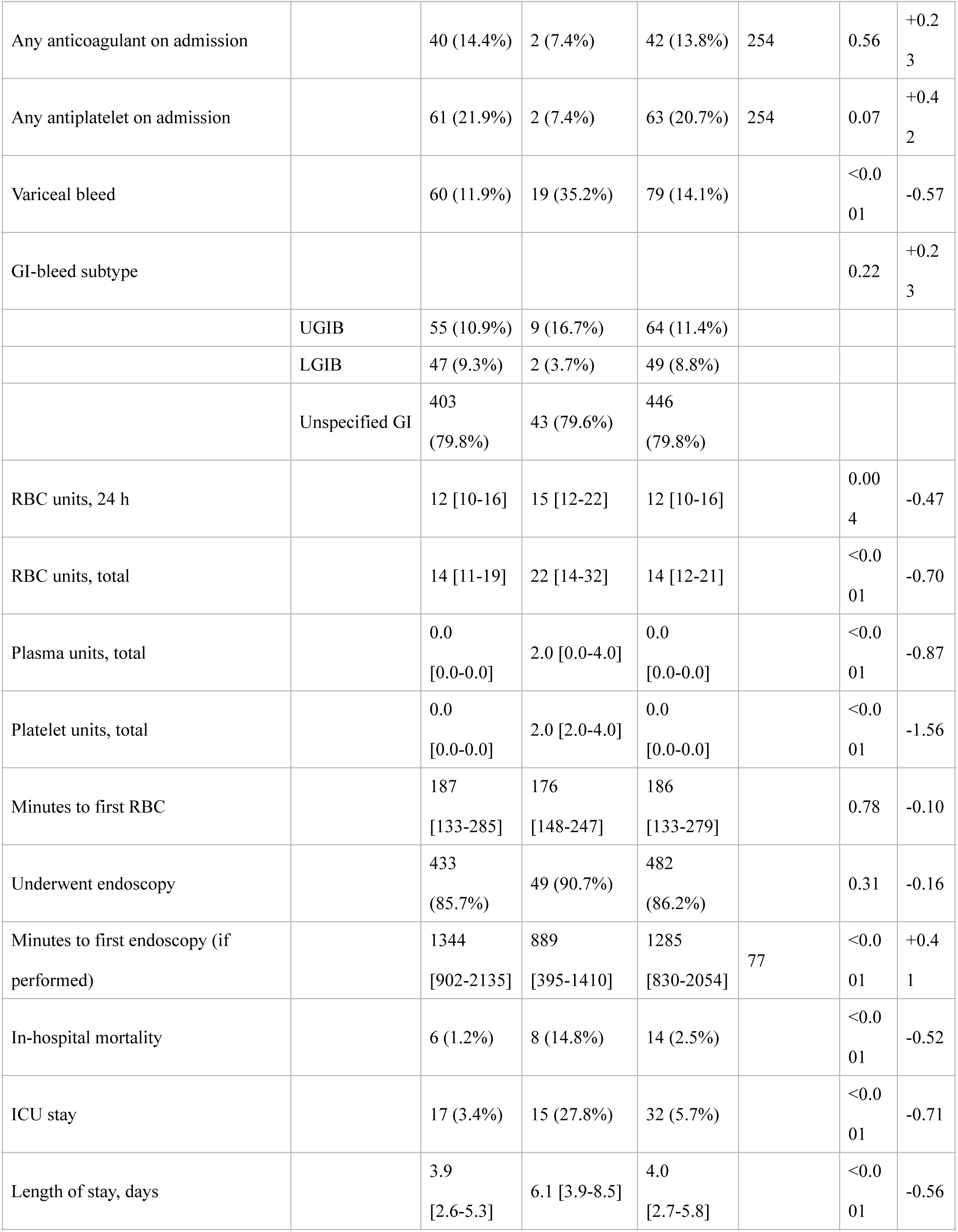
Baseline characteristics and outcomes by transfusion exposure. Baseline demographics, admission laboratories, transfusion quantities, and unadjusted outcomes stratified by exposure. Continuous variables reported as median [Q1–Q3]; categorical as n (%). SMD = standardised mean difference (RBC-only − Balanced).

### Propensity-score model and covariate balance

The propensity-score model was fit on the complete-case set for the 10 pre-specified covariates (10 encounters dropped: 2 for missing platelet count in 24 hours and 8 for missing BUN in 24 hours; analytic n = 549) (Figure 1). Figure 2 shows that several baseline covariates had |SMD| > 0.30 between arms, including age, first platelet count, variceal bleed, and number of RBC units within 24 hours. After IPTW, binary covariates achieved balance (|SMD| < 0.10 for endoscopy and variceal bleed), continuous and multicategory covariates retained residual imbalance in the 0.14–0.29 range (largest: bleeding subtype 0.29; first haemoglobin level 0.28; first platelet count 0.21). This residual imbalance is a consequence of the small effective sample size on the Balanced arm (Kish effective n ≈ 30). The distribution of IPTW weights (Figure 3) was skewed but within expected bounds after one-sided winsorisation at the 99th percentile.

**Figure 1.**
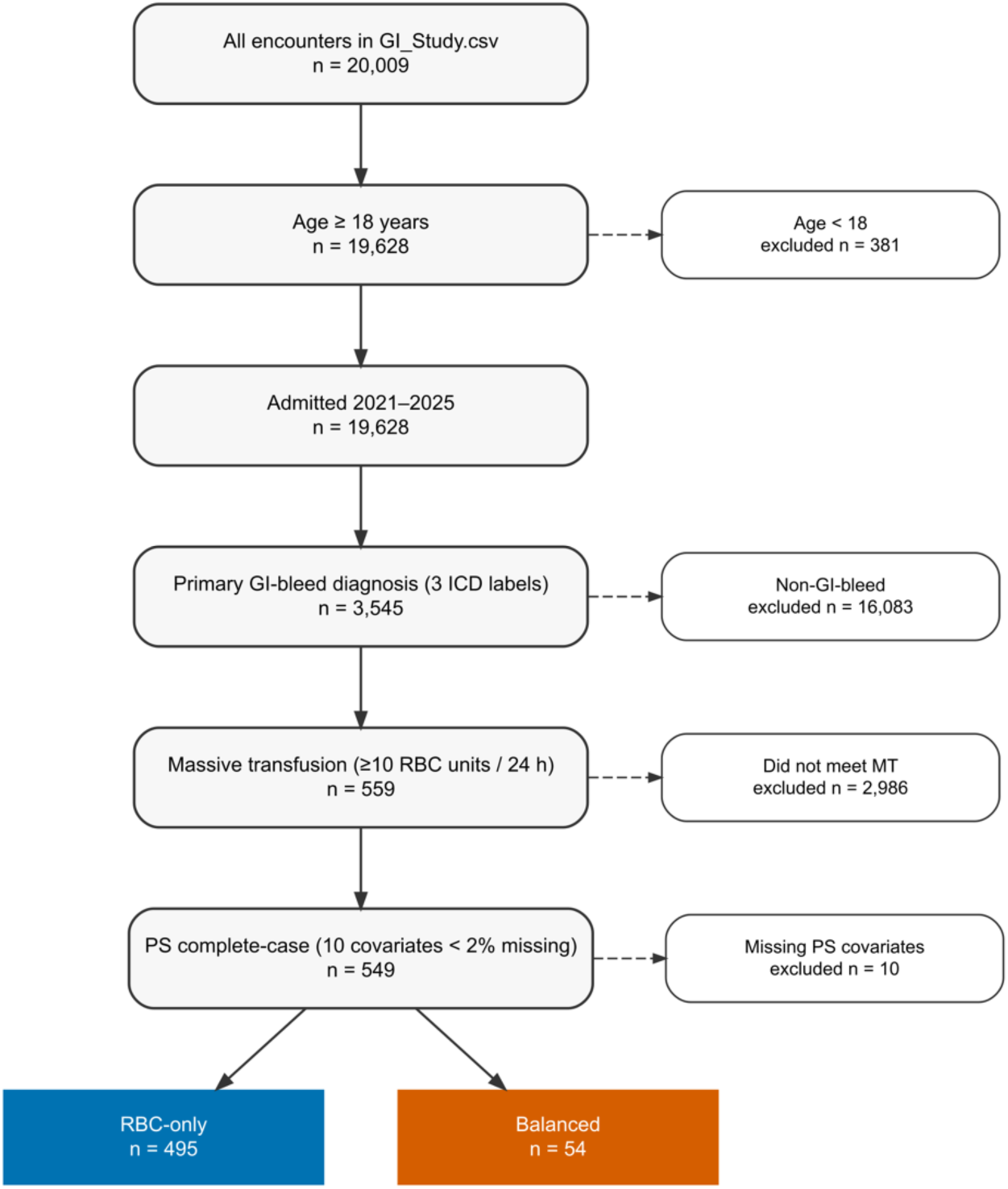
CONSORT-style inclusion cascade. Flow from the full 2021–2025 inpatient extract through age, diagnosis, and massive-transfusion filters to the analytic cohort of 559 encounters in 536 patients. See Methods (Cohort definition).

**Figure 2.**
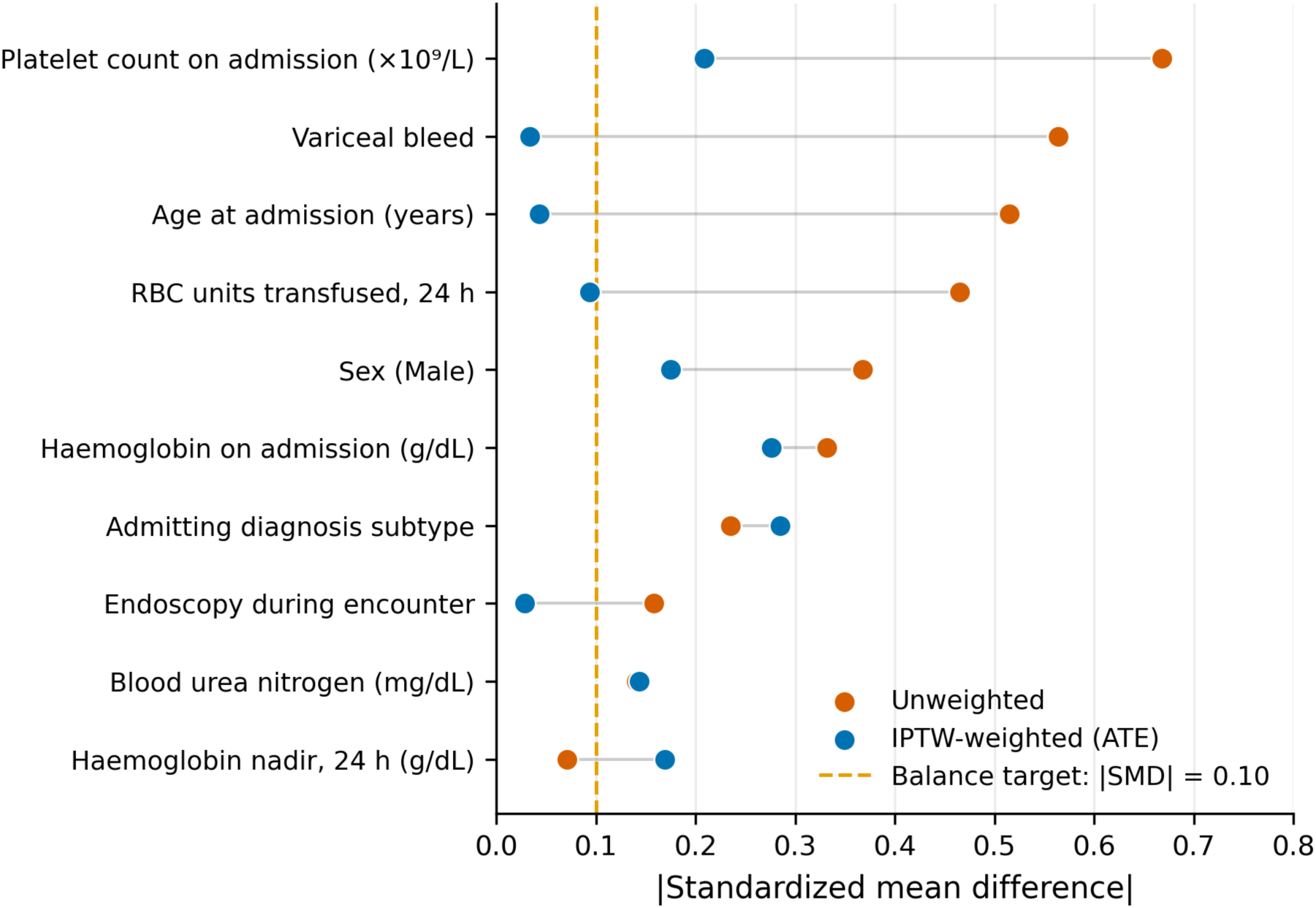
Love plot of covariate balance pre- and post-IPTW. Standardised mean differences (SMD) between Balanced and RBC-only arms for the 10 pre-specified propensity-score covariates, before (open circles) and after (filled circles) inverse-probability-of-treatment weighting. Dashed vertical lines indicate the ±0.10 balance threshold.

**Figure 3.**
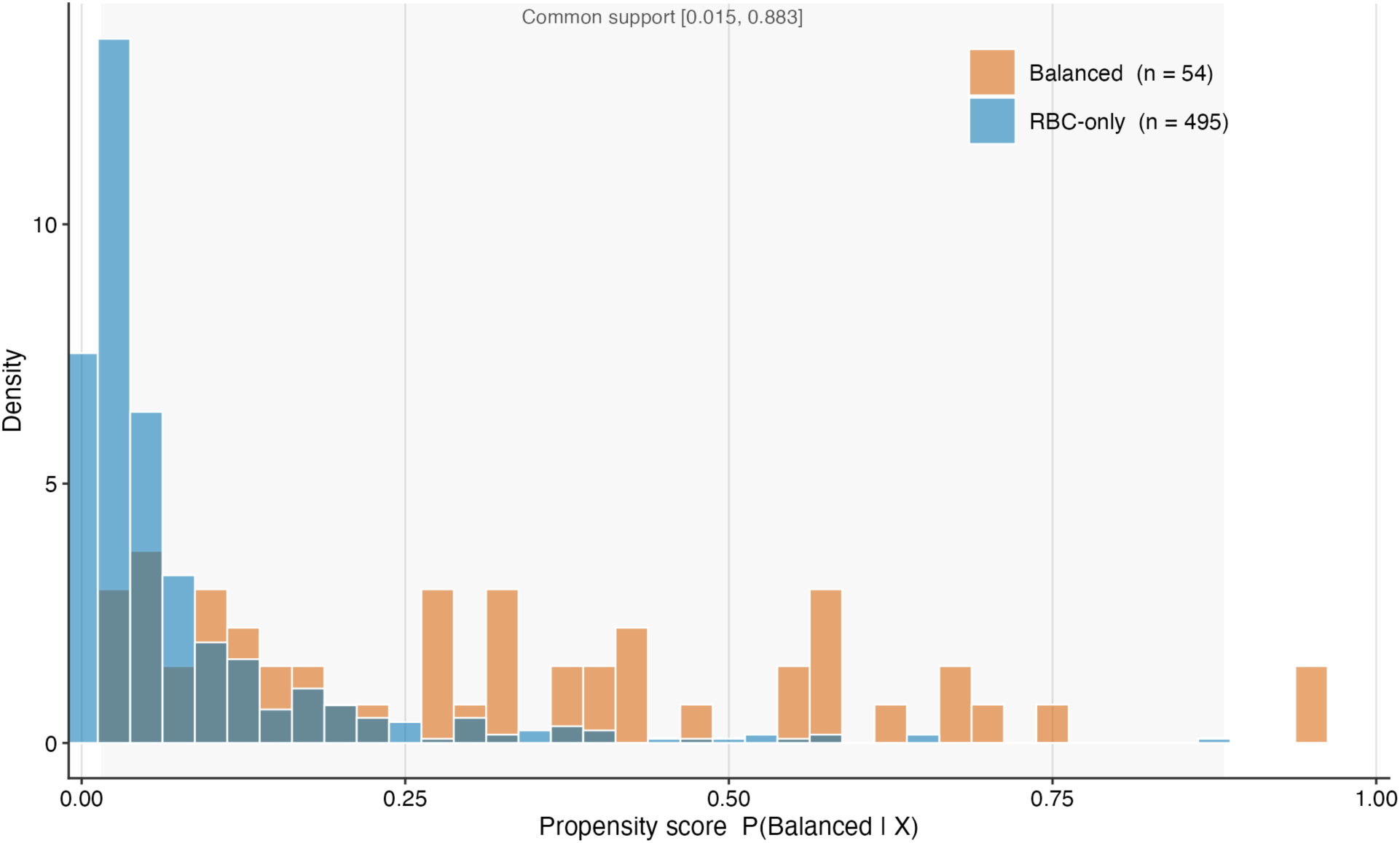
Distribution of IPTW weights by exposure arm. Box-plot and stripplot of IPTW weights after one-sided winsorisation at the 99th percentile. The Balanced arm carries higher, more variable weights, consistent with propensity-score overlap but limited effective sample size.

### Primary outcome: in-hospital mortality

The AIPTW-estimated in-hospital mortality risk was 21.0% under Balanced and 1.2% under RBC-only (Table 2). The adjusted risk difference (RBC-only − Balanced) was -19.8 pp (95% CI -68.1 to -2.2 pp; upper 97.5% confidence bound -2.2 pp). The upper 97.5% bound lies below both the pre-specified primary margin of +5 pp and the more stringent sensitivity margin of +3 pp; non-inferiority is therefore demonstrated at both margins. The point estimate is directionally consistent with potential superiority of the RBC-only strategy, although this is not formally tested.

**Table 2.** Primary and sensitivity estimates for the risk difference in in-hospital mortality. AIPTW risk differences (RBC-only − Balanced) with 95% confidence intervals and the upper 97.5% non-inferiority bound. The pre-specified primary margin is 5 pp; the sensitivity margin is 3 pp. The primary analysis and all sensitivities demonstrate non-inferiority at the primary margin.

| Analysis | n | RD (pp) | 95% CI (pp) | Upper 97.5% (pp) |
| --- | --- | --- | --- | --- |
| Primary AIPTW | 549 | -19.8 | -68.1 to -2.2 | -2.2 |
| Sensitivity 1: 1:1 PS matching | 98 | -14.3 | -24.5 to -6.1 | -6.1 |
| Sensitivity 2: complete-case w/ INR | 454 | -18.0 | -52.9 to -1.7 | -1.7 |
| Sensitivity 3: broader GI diagnosis | 749 | -17.2 | -42.8 to -1.3 | -1.3 |
| Sensitivity 4: first encounter per | 527 | -22.1 | -90.2 to -1.8 | -1.8 |

*E-value (Methods sensitivity 5): 35.7 for the point estimate, 4.80 for the upper 97.5% bound.*

### Secondary outcomes

The adjusted risk difference for ICU admission (RBC-only − Balanced) was -17.3 pp (95% CI -55.2 to +0.7 pp), directionally consistent with the mortality result but not formally tested for non-inferiority. The Hodges–Lehmann median difference in LOS (RBC-only − Balanced) was -2.0 days (95% CI -3.5 to -0.9 days, p=0.001).

### Sensitivity analyses

All pre-specified sensitivity analyses were concordant with the primary result on direction and magnitude (Figure 4; Table 2). One-to-one propensity-score matching (n = 98 in 49 pairs) yielded an RD of −14.3 pp (95% CI −24.5 to −6.1 pp). Complete-case AIPTW including INR (n = 454) gave an RD of −18.0 pp (95% CI −52.9 to −1.7 pp). Broadening the admitting-diagnosis inclusion via a text-based regular expression that captured similar named GI-bleed entities (n = 749; +200 encounters relative to the primary cohort) gave −17.2 pp (95% CI −42.8 to −1.3 pp), with the upper 97.5% bound (−1.3 pp) below both pre-specified margins. Restriction to the first MT encounter per patient (n = 527) gave −22.1 pp (95% CI −90.2 to −1.8 pp). The E-value for the point estimate was 35.7; the E-value for the upper 97.5% bound was 4.80.

**Figure 4.**
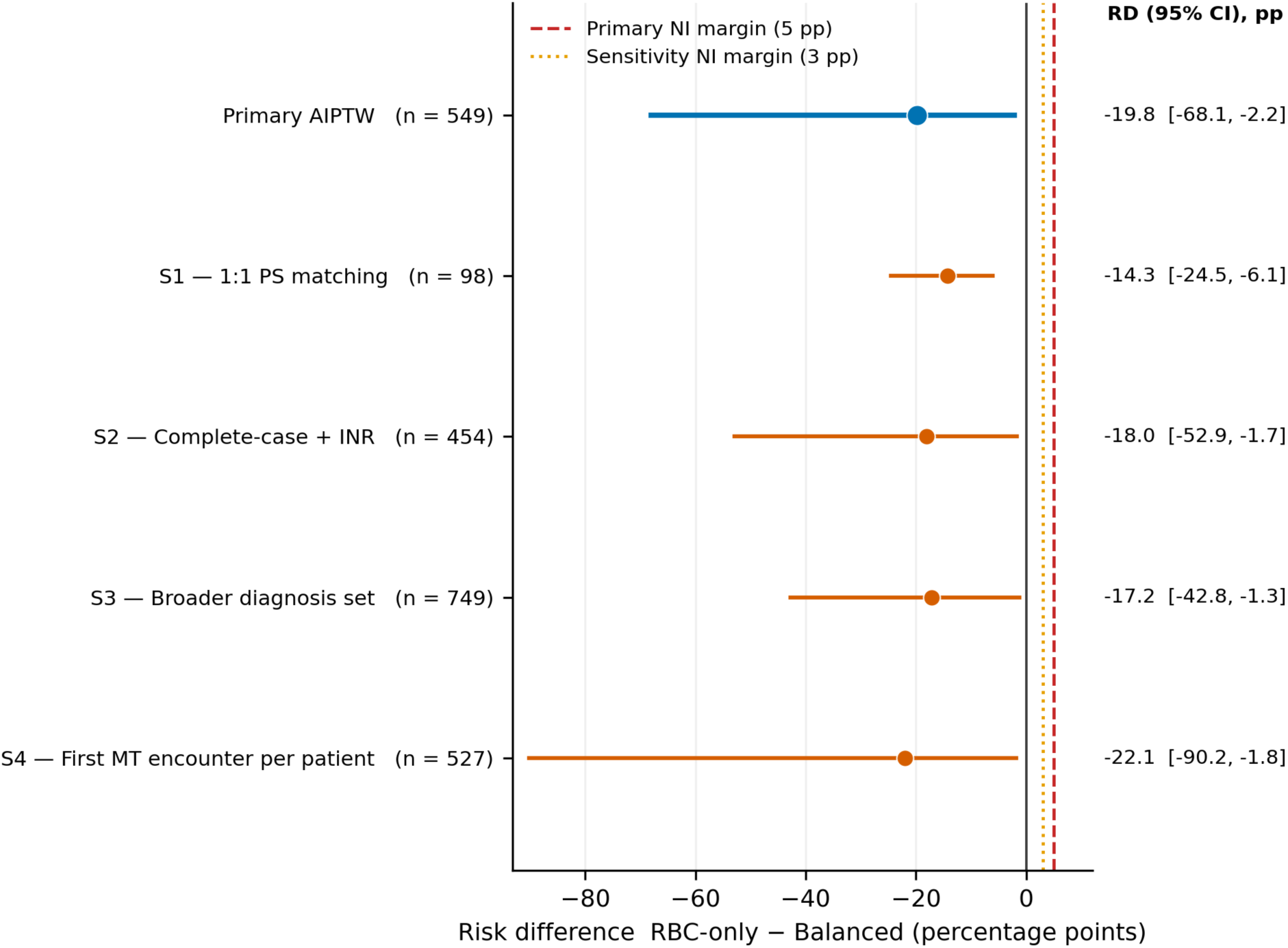
Forest plot of primary and sensitivity estimates. AIPTW risk differences (RBC-only − Balanced) for in-hospital mortality, with 95% confidence intervals. Vertical reference lines mark the pre-specified 5-pp primary and 3-pp sensitivity non-inferiority margins. Five analyses (Primary, S1, S2, S3, and S4) demonstrate non-inferiority at the 5-pp primary margin. The E-value sensitivity (Methods item 5) is not a risk difference and is reported separately in Table 2.

## Discussion

In a propensity-score–weighted cohort of adults meeting massive-transfusion criteria for gastrointestinal haemorrhage, an RBC-only transfusion strategy was non-inferior to a balanced (RBC + plasma and/or platelets) strategy for in-hospital mortality at both the pre-specified 5-pp margin and a more stringent 3-pp sensitivity margin. The finding was robust across five pre-specified sensitivity analyses, including propensity-score matching, a complete-case model incorporating admission INR, a broader text-based diagnosis inclusion, a first-encounter restriction, and an E-value sensitivity for unmeasured confounding.

These results address a longstanding ambiguity in non-trauma massive haemorrhage: whether the strategy of damage control resuscitation (DCR), an empirical, fixed-ratio plasma and platelet support derived from trauma physiology, must accompany the operational architecture of a massive transfusion protocol (MTP) when activation occurs in a GI-bleed patient. The two are conceptually distinct. DCR is the resuscitation strategy that prescribes pre-emptive ratio-based component therapy, permissive hypotension, and early haemorrhage control, predicated on the physiology of acute traumatic coagulopathy.^6, 8^ MTP is the delivery system, an institutional trigger, the pre-thawed plasma, the fixed-ratio cooler that makes rapid component access feasible.^7^ In trauma the two are bundled, and the PROPPR trial established this bundle as the practice standard.^9^ Their transfer into non-trauma haemorrhage has been largely empirical rather than physiologically derived. The rationale for DCR ratios in trauma is to manage acute traumatic coagulopathy driven by tissue injury, shock, and hyperfibrinolysis,^14, 15, 32^ which is not the state of most patients with GI haemorrhage, whose coagulation abnormalities are more commonly sequelae of chronic liver disease, anticoagulant or antiplatelet therapy, or dilutional effects of large-volume RBC resuscitation.^13, 16, 17, 19^ Empirical component support in this physiology therefore exposes patients to the known risks of allogeneic plasma and platelets with transfusion-associated circulatory overload, transfusion-related acute lung injury, febrile and allergic reactions, and rare infectious transmission^20, 33^ without a mechanistic basis for a compensating benefit. Our findings support decoupling the MTP delivery architecture from obligatory DCR ratios in massive GI haemorrhage, in favour of individualised, physiologically guided component use.

Three features distinguish the present analysis from prior literature on transfusion strategy in GI haemorrhage. First, earlier non-trauma massive-transfusion analyses examined plasma:RBC ratios but did not directly test the policy-relevant question of whether plasma and platelets can be safely withheld altogether; we frame the question as non-inferiority of RBC-only versus any balanced exposure.^12, 13^ Second, we use a doubly robust AIPTW estimator with a pre-specified 10-covariate propensity model and five sensitivity analyses, reducing dependence on any single modelling choice. Third, the cohort is restricted to encounters meeting strict massive-transfusion criteria (≥ 10 RBC units within 24 hours), aligning the comparison with the operational threshold at which institutional MTP-style resuscitation is actually triggered, rather than with smaller-volume bleeds in which component decisions are physiologically and clinically distinct.

Our results complement and extend the prior literature on transfusion thresholds in GI haemorrhage. Villanueva and colleagues demonstrated that a restrictive RBC transfusion threshold (haemoglobin 7 g/dL) reduced 45-day mortality compared with a liberal threshold (9 g/dL) in patients with acute upper GI bleeding;^2^ TRIGGER confirmed the feasibility of this approach at scale.^1^ Our analysis addresses a distinct axis of the transfusion-strategy question: whether component composition — rather than RBC transfusion threshold — independently determines outcomes in massive GI haemorrhage. Taken together with the restrictive-threshold evidence, the data suggest that GI haemorrhage responds better to targeted, physiologically guided component use than to protocolised 1:1:1 resuscitation.

The HALT-IT trial provides a close conceptual analogue for our findings. Tranexamic acid, a antifibrinolytic agent that reduces mortality in trauma patients with significant bleeding,^34^ was tested in 12,009 patients with acute GI bleeding and conferred no mortality reduction while increasing venous thromboembolism and seizures.^3^ Haemostatic interventions that rescue coagulopathic trauma patients from exsanguination must be tested, rather than assumed to benefit a GI-bleed population whose acute coagulopathy is qualitatively different.

The cohort’s substantial representation of variceal bleeding (11.9% in the RBC-only arm and 35.2% in the Balanced arm) adds another important point to the case for individualised component use. In portal hypertension, aggressive volume expansion, particularly with plasma, can raise portal venous pressure and precipitate rebleeding, and the Baveno VII consensus accordingly cautions against over-transfusion of any component in this subgroup.^18^ A default 1:1:1 protocol, if applied without attention to the underlying physiology, risks converting a presumed haemostatic intervention into a haemodynamic injury.

### Limitations

Several limitations merit emphasis. First, plasma and platelet count in the source extract are whole-encounter totals, not 24-hour totals; the RBC:plasma:platelet ratios conventionally reported in trauma MT literature could not therefore be computed on the acute resuscitation window. We assumed that patients are more likely to receive plasma and platelets during a massive bleed event which would be early in the admission, rather than later in the hospital stay downstream. In our cohort 67% of patients received their entire RBC load within the first 24 hours and 85% of patients received ≥ 75% of total transfusions in 24 hours, so encounter totals approximate acute transfusion. This pattern suggests that a fraction of the plasma and platelet units attributed to Balanced were given outside the acute resuscitation window (*e.g.*, peri-endoscopic platelet support or delayed plasma for anticoagulant reversal).

Second, the 9:1 exposure imbalance constrains precision. The Balanced arm contains only 54 encounters and 8 in-hospital deaths and effective small sample size (Kish ESS ≈ 30). Pre-specified power calculations showed that, at the observed baseline mortality of approximately 15% in the Balanced arm, the 5-pp primary margin has adequate power (≥ 80%). The present study is therefore well-positioned to rule out large harms (≥ 10 pp) of an RBC-only strategy but would be inconclusive against tighter margins. The residual imbalance on continuous covariates after weighting (|SMD| 0.14–0.29 on six of ten covariates) is also caused by the small effective Balanced-arm sample. The doubly-robust AIPTW estimator is utilised to augment the residual continuous imbalance.

Third, admission creatinine could not be used as a propensity-score covariate due to non-serum sample contamination identified in the source data. In addition, baseline platelet count differed substantially between arms (median 140 × 10⁹/L Balanced vs 229 × 10⁹/L RBC-only) and was among the covariates retaining residual imbalance after weighting (|SMD| 0.21); first platelet count is explicitly included in the AIPTW outcome regression to limit confounding by this clinical predictor.

Forth, this is a single-centre retrospective analysis of structured electronic health record, and unmeasured confounding cannot be excluded. The E-value of 4.80 for the upper 97.5% bound indicates that a hypothetical unmeasured confounder would need to be associated with both exposure and outcome by a risk ratio of that magnitude, beyond the measured covariates, to overturn the non-inferiority conclusion; such a confounder is doubtful given the richness of the adjustment set.

Fifth, the dataset captures the total number of RBC units transfused within 24 hours but not their temporal distribution. Rapid, sustained haemorrhage requiring 8 units over 6 hours generates substantially greater acute dilution of coagulation factors than an equivalent total delivered in interrupted episodes during which clotting factors may partially recover, whereas dilutional thrombocytopenia is less rate-sensitive and reflects cumulative platelet loss across the resuscitation. This rate-and-pattern asymmetry was not captured in the source extract and could independently influence both the clinical decision to administer plasma or platelets and downstream patient outcomes. Future prospective studies should capture the timing and rate of component delivery.

Finally, the cohort reflects practice at a single institution between 2021 and 2025 and may not generalise to settings with systematically different MT-protocol triggers, case mix, or endoscopic capacity. Prospective multi-centre confirmation is warranted.

### Conclusions

In this propensity-score–weighted cohort of adults with massive GI haemorrhage, an RBC-only transfusion strategy was non-inferior to a balanced strategy for in-hospital mortality at both 5-pp and 3-pp margins. The findings support managing acute large-volume GI haemorrhage with adequate RBC transfusion as the primary resuscitative component, reserving plasma and platelets for patients with ongoing bleeding requiring sustained transfusion (*e.g.*, ≥ 1 unit RBC every 1–2 hours for 6–8 hours). Prospective confirmation is warranted.

## Supporting information

power calculations

STROBE checklist

CONSORT-NI extension

## Statements

### Contributors

BB conceived the study, designed the analysis plan, curated the dataset, conducted the statistical analyses in parallel R and Python implementations, and drafted the manuscript. JDS contributed to the conception of the study, provided clinical interpretation of the cohort, reviewed and revised the manuscript. CN contributed to the conception of the study, provided clinical interpretation of the cohort, reviewed and revised the manuscript. contributed to the conception of the study and revised the manuscript. BB is the guarantor and accepts full responsibility for the conduct of the study and has access to the data. All listed authors approved the final version.

### Ethics approval

This study was approved under umbrella protocol 2339166-1, “Design and implementation of a transfusion medicine data science platform” (CMTT-PROJ 210925; effective 2025-10-01) as a pre-specified sub-analysis; no separate IRB submission was required. The requirement for individual patient consent was waived given the retrospective use of de-identified electronic-health-record data.

### Data availability statement

The underlying electronic-health-record data contain potentially identifiable protected health information and cannot be publicly shared. De-identified analytic extracts sufficient to reproduce the primary and sensitivity estimates may be made available to qualified investigators upon reasonable request to the corresponding author and subject to institutional data-use agreement and IRB approval. All analytic code (parallel R and Python implementations) will be deposited in an open repository on acceptance.

## Ethics

Approved under umbrella protocol 2339166-1 (“Design and implementation of a transfusion medicine data science platform”; CMTT-PROJ 210925; effective 2025-10-01). This is a pre-specified sub-analysis; no separate IRB submission was required.

## References

1. Jairath V, Kahan BC, Gray A. Restrictive versus liberal blood transfusion for acute upper gastrointestinal bleeding (TRIGGER): a pragmatic, open-label, cluster randomised feasibility trial. Lancet 2015;386(9989):137–44.

2. Villanueva C, Colomo A, Bosch A. Transfusion strategies for acute upper gastrointestinal bleeding. N Engl J Med 2013;368(1):11–21.

3. Halt-It Trial Collaborators. Effects of a high-dose 24-h infusion of tranexamic acid on death and thromboembolic events in patients with acute gastrointestinal bleeding (HALT-IT): an international randomised, double-blind, placebo-controlled trial. Lancet 2020;395(10241):1927–36.

4. Harris T, Davenport R, Mak M, et al. The Evolving Science of Trauma Resuscitation. Emergency Medicine Clinics of North America 2018;36(1):85–106. doi: 10.1016/j.emc.2017.08.009

5. Hooper N, Armstrong TJ. Hemorrhagic Shock. StatPearls. Treasure Island (FL)2026.

6. Walsh M, Moore EE, Moore HB, et al. Whole Blood, Fixed Ratio, or Goal-Directed Blood Component Therapy for the Initial Resuscitation of Severely Hemorrhaging Trauma Patients: A Narrative Review. Journal of Clinical Medicine 2021; 10(2).

7. Curry N, Stanworth S, Hopewell S, et al. Trauma-Induced Coagulopathy – A Review of the Systematic Reviews: Is There Sufficient Evidence to Guide Clinical Transfusion Practice? Transfusion Medicine Reviews 2011;25(3):217–31.

8. Ntourakis D, Liasis L. Damage control resuscitation in patients with major trauma: prospects and challenges. Journal of Emergency and Critical Care Medicine 2020;4

9. Holcomb JB, Tilley BC, Baraniuk S. Transfusion of plasma, platelets, and red blood cells in a 1:1:1 vs a 1:1:2 ratio and mortality in patients with severe trauma: the PROPPR randomized clinical trial. JAMA 2015;313(5):471–82.

10. Duchesne JC, Hunt JP, Wahl G, et al. Review of current blood transfusions strategies in a mature level I trauma center: were we wrong for the last 60 years? *The Journal of Trauma: Injury*, Infection, and Critical Care 2008;65(2):272–78. doi: 10.1097/TA.0b013e31817e5166

11. Odutayo A, Desborough MJR, Trivella M, et al. Restrictive versus liberal blood transfusion for gastrointestinal bleeding: a systematic review and meta-analysis of randomised controlled trials. The Lancet Gastroenterology & Hepatology 2017;2(5):354–60. doi: 10.1016/S2468-1253(17)30054-7

12. Mesar T, Larentzakis A, Dzik W, et al. Association Between Ratio of Fresh Frozen Plasma to Red Blood Cells During Massive Transfusion and Survival Among Patients Without Traumatic Injury. JAMA Surg 2017;152(6):574–80. doi: 10.1001/jamasurg.2017.0098

13. Etchill EW, Myers SP, McDaniel LM, et al. Should All Massively Transfused Patients Be Treated Equally? An Analysis of Massive Transfusion Ratios in the Nontrauma Setting. Crit Care Med 2017;45(8):1311–16. doi: 10.1097/CCM.0000000000002498

14. Brohi K, Singh J, Heron M, et al. Acute traumatic coagulopathy. The Journal of Trauma: Injury, Infection, and Critical Care 2003;54(6):1127–30. doi: 10.1097/01.TA.0000069184.82147.06

15. Cardenas JC, Wade CE, Holcomb JB. Mechanisms of trauma-induced coagulopathy. Curr Opin Hematol 2014;21(5):404–09.

16. Sengupta N, Feuerstein JD, Jairath V. Management of patients with acute lower gastrointestinal bleeding: an updated ACG guideline. Am J Gastroenterol 2023;118(2):208–31.

17. Laine L, Barkun AN, Saltzman JR, et al. ACG Clinical Guideline: Upper Gastrointestinal and Ulcer Bleeding. Am J Gastroenterol 2021;116(5):899–917. doi: 10.14309/ajg.0000000000001245

18. de Franchis R, Bosch J, Garcia-Tsao G. Baveno VII — Renewing consensus in portal hypertension. J Hepatol 2022;76(4):959–74.

19. Siau K, Hearnshaw S, Stanley AJ, et al. British Society of Gastroenterology (BSG)-led multisociety consensus care bundle for the early clinical management of acute upper gastrointestinal bleeding. Frontline Gastroenterology 2020;11(4):311. doi: 10.1136/flgastro-2019-101395

20. Bulle EB, Klanderman RB, Pendergrast J, et al. The recipe for TACO: A narrative review on the pathophysiology and potential mitigation strategies of transfusion-associated circulatory overload. Blood reviews 2022;52:100891.

21. Fabricius R, Svenningsen P, Hillingso J, et al. Effect of Transfusion Strategy in Acute Non-variceal Upper Gastrointestinal Bleeding: A Nationwide Study of 5861 Hospital Admissions in Denmark. World J Surg 2016;40(5):1129–36. doi: 10.1007/s00268-015-3370-4

22. Hébert PC, Wells G, Blajchman MA. A multicenter, randomized, controlled clinical trial of transfusion requirements in critical care. N Engl J Med 1999;340(6):409–17.

23. Holst LB, Haase N, Wetterslev J. Lower versus higher hemoglobin threshold for transfusion in septic shock. N Engl J Med 2014;371(15):1381–91.

24. Austin PC. An introduction to propensity score methods for reducing the effects of confounding in observational studies. Multivariate Behav Res 2011;46(3):399–424.

25. Kang JDY, Schafer JL. Demystifying Double Robustness: A Comparison of Alternative Strategies for Estimating a Population Mean from Incomplete Data. Statistical Science 2007;22(4):523–39.

26. Lunceford JK, Davidian M. Stratification and weighting via the propensity score in estimation of causal treatment effects: a comparative study. Stat Med 2004;23(19):2937–60.

27. von Elm E, Altman DG, Egger M. The Strengthening the Reporting of Observational Studies in Epidemiology (STROBE) statement: guidelines for reporting observational studies. PLoS Med 2007;4(10):e296.

28. Piaggio G, Elbourne DR, Altman DG, et al. Reporting of noninferiority and equivalence randomized trials: extension of the CONSORT 2010 statement. JAMA 2012;308(24):2594–604. doi: 10.1001/ jama.2012.87802

29. Funk MJ, Westreich D, Wiesen C, et al. Doubly robust estimation of causal effects. American journal of epidemiology 2011;173(7):761–67. doi: 10.1093/aje/kwq439

30. Austin PC. Balance diagnostics for comparing the distribution of baseline covariates between treatment groups in propensity-score matched samples. Stat Med 2009;28(25):3083–107.

31. VanderWeele TJ, Ding P. Sensitivity analysis in observational research: introducing the E-value. Ann Intern Med 2017;167(4):268–74.

32. Wada H, Matsumoto T, Yamashita Y. Diagnosis and treatment of disseminated intravascular coagulation (DIC) according to four DIC guidelines. J Intensive Care 2014;2(1):15.

33. Semple JW, Rebetz J, Kapur R. Transfusion-associated circulatory overload and transfusion-related acute lung injury. Blood 2019;133(17):1840–53. doi: 10.1182/blood-2018-10-860809 [published Online First: 20190226]

34. Crash- trial collaborators. Effects of tranexamic acid on death, vascular occlusive events, and blood transfusion in trauma patients with significant haemorrhage (CRASH-2): a randomised, placebo-controlled trial. Lancet 2010;376(9734):23–32. doi: 10.1016/S0140-6736(10)60835-5 [published Online First: 20100614]

