## Supplementary material for "Non-inferiority of a red-blood-cell–only transfusion strategy compared with balanced resuscitation in adults with massive gastrointestinal haemorrhage: a propensity-score–weighted cohort study": power calculations

### Supplement S-Power: Feasibility of the non-inferiority analysis

Power of the one-sided (α = 0.025) non-inferiority test for the risk difference in in-hospital mortality, under the observed arm sizes (n_RBC-only = 495, n_balanced = 54; propensity-score complete-case). Values are from the asymptotic normal approximation to the unadjusted RD Z-test and represent a first-order envelope on the AIPTW estimator's precision; after IPTW, the effective sample sizes shrink to Kish-ESS of 450.9 (RBC-only) and 29.6 (balanced) because of the concentration of weight on the small balanced arm.

#### Power at the primary 5-pp margin (raw arm sizes)

| | p0=5% | p0=10% | p0=15% | p0=20% |
|:---|---:|---:|---:|---:|
| -15.0 pp | nan | nan | nan | 0.951 |
| -10.0 pp | nan | nan | 0.857 | 0.763 |
| -5.0 pp | nan | 0.663 | 0.509 | 0.422 |
| -3.0 pp | 0.751 | 0.471 | 0.351 | 0.289 |
| +0.0 pp | 0.360 | 0.213 | 0.163 | 0.138 |
| +2.0 pp | 0.155 | 0.102 | 0.084 | 0.075 |

#### Power at the sensitivity 3-pp margin (raw arm sizes)

| | p0=5% | p0=10% | p0=15% | p0=20% |
|:---|---:|---:|---:|---:|
| -15.0 pp | nan | nan | nan | 0.902 |
| -10.0 pp | nan | nan | 0.746 | 0.640 |
| -5.0 pp | nan | 0.478 | 0.354 | 0.291 |
| -3.0 pp | 0.508 | 0.293 | 0.218 | 0.182 |
| +0.0 pp | 0.159 | 0.103 | 0.085 | 0.075 |
| +2.0 pp | 0.050 | 0.042 | 0.039 | 0.037 |

#### Power at 5-pp margin using Kish effective N (IPTW-adjusted)

| | p0=5% | p0=10% | p0=15% | p0=20% |
|:---|---:|---:|---:|---:|
| -15.0 pp | nan | nan | nan | 0.768 |
| -10.0 pp | nan | nan | 0.617 | 0.517 |
| -5.0 pp | nan | 0.429 | 0.319 | 0.263 |
| -3.0 pp | 0.504 | 0.293 | 0.220 | 0.183 |
| +0.0 pp | 0.226 | 0.140 | 0.111 | 0.097 |
| +2.0 pp | 0.107 | 0.075 | 0.064 | 0.059 |

#### Minimum detectable NI margin at 80% and 90% power

Margin δ (pp) needed to declare non-inferiority under the specified true RD and baseline mortality. True RD = 0 corresponds to the null of identical mortality in the two strategies.

##### raw (unweighted)

**True RD = −3 pp**

| | p0=5% | p0=10% | p0=15% | p0=20% |
|:---|---:|---:|---:|---:|
| 80% power | 5.5 | 8.9 | 11.2 | 13 |
| 90% power | 6.8 | 10.7 | 13.4 | 15.5 |

**True RD = +0 pp**

| | p0=5% | p0=10% | p0=15% | p0=20% |
|:---|---:|---:|---:|---:|
| 80% power | 8.8 | 12 | 14.3 | 16.1 |
| 90% power | 10.1 | 13.9 | 16.6 | 18.6 |

##### Kish ESS (IPTW)

**True RD = −3 pp**

| | p0=5% | p0=10% | p0=15% | p0=20% |
|:---|---:|---:|---:|---:|
| 80% power | 8.4 | 12.8 | 15.9 | 18.2 |
| 90% power | 10.2 | 15.3 | 18.9 | 21.5 |

**True RD = +0 pp**

| | p0=5% | p0=10% | p0=15% | p0=20% |
|:---|---:|---:|---:|---:|
| 80% power | 11.6 | 16 | 19 | 21.3 |
| 90% power | 13.4 | 18.5 | 22 | 24.6 |

#### Interpretation

Given the 9:1 exposure imbalance, the balanced arm is the binding constraint on precision. At the observed baseline mortality rate of ~15% in the balanced arm, the 5-pp primary margin has adequate power (≥ 80%) only when the true RD is at or below ~0 pp in favour of RBC-only; if the two strategies are truly equivalent, the minimum detectable margin at 80% power using the raw arm sizes is 14.3 pp, widening to 19.0 pp when the IPTW effective sample size is used. The study is therefore well-positioned to rule out large harms (≥ 10 pp) but will be inconclusive against tighter margins unless the true RD is clearly negative. These limits are acknowledged in the Discussion and motivate presenting the upper 97.5% confidence bound alongside the point estimate for every analysis.

#### Caveats

1. Formulas use the asymptotic normal approximation to the unadjusted

RD test; with only 8 deaths in the balanced arm, small-sample deviations exist. The bootstrap percentile CI used for the primary inference is preferred and may be slightly wider.

2. Kish's effective sample size is a summary of weight concentration, not a full replacement for the IPTW design effect; it understates precision for the AIPTW estimator, which borrows strength from the outcome regression.

3. True-RD scenarios should be read relative to the primary conservative intent: the study is powered to demonstrate NI only if RBC-only is at least as safe as balanced; it is not powered to establish superiority.
