## Supplementary material for "Non-inferiority of a red-blood-cell–only transfusion strategy compared with balanced resuscitation in adults with massive gastrointestinal haemorrhage: a propensity-score–weighted cohort study": STROBE checklist

### STROBE Statement

| Item No. | Topic | STROBE recommendation | Response | Page / Line |
|---|---|---|---|---|
| **Title and abstract** | | | | |
| **1 (a)** | **Title and abstract** | Indicate the study's design with a commonly used term in the title or the abstract. | Retrospective single-center cohort study; identified as in the Design field of the structured abstract. Non-inferiority framing is stated explicitly in title and abstract. | p.1, L1–2 (title); p.2, L9 (abstract Design) |
| **1 (b)** | | Provide in the abstract an informative and balanced summary of what was done and what was found. | Structured abstract reports cohort definition, exposure (RBC-only vs Balanced), primary outcome (in-hospital mortality), AIPTW estimator, pre-specified NI margin (5 percentage points absolute risk difference, one-sided α=0.025), and the upper 97.5% confidence bound relative to that margin. The Significance of this study box covers what is already known, what this study adds, and how this might affect research/practice/policy. | p.2, L1–25 |
| **Introduction** | | | | |
| **2** | **Background / rationale** | Explain the scientific background and rationale for the investigation being reported. | Trauma requires a balanced 1:1:1 resuscitation (PROPPR; Holcomb 2015) contrasts with the physiology of massive GI hemorrhage, where the trauma triad (coagulopathy, hypothermia, acidosis) is rare (HALT-IT, Baveno VII, TRIGGER, TRICC, TRISS). | p.4, L1–27; p.5, L1–14 |
| **3** | **Objectives** | State specific objectives, including any prespecified hypotheses. | Prespecified objective: to test non-inferiority of an RBC-only versus a Balanced transfusion strategy with respect to in-hospital mortality in adults requiring massive transfusion for GI hemorrhage. Primary NI margin 5 pp absolute RD (one-sided α=0.025); pre-specified sensitivity margin 3 pp on average treatment effect (ATE). | p.5, L15–21 |
| **Methods** | | | | |

| Item No. | Topic | STROBE recommendation | Response | Page / Line |
|---|---|---|---|---|
| 4 | **Study design** | Present key elements of study design early in the paper. | Retrospective cohort study with a non-inferiority hypothesis. Confounding addressed by augmented inverse-probability-of-treatment weighting (AIPTW; doubly robust). Reporting borrows CONSORT-NI discipline; STROBE used as primary checklist. | p.6, L2–11 |
| 5 | **Setting** | Describe the setting, locations, and relevant dates, including periods of recruitment, exposure, follow-up, and data collection. | Single tertiary-care center. Eligible admissions 1 January 2021 through 31 December 2025 (60-month window). Data extracted from the institutional transfusion-medicine data science platform (IRB #2339166-1; umbrella protocol "Design and implementation of a transfusion medicine data science platform," effective 1 October 2025). Follow-up is in-hospital only. | p.6, L2–11 |
| 6 (a) | **Participants** | Give the eligibility criteria, and the sources and methods of selection of participants. Describe methods of follow-up. | Inclusion: adult (≥18 y), inpatient encounter; with primary admitting diagnosis of Upper GI Hemorrhage, Lower GI Hemorrhage, Gastrointestinal Hemorrhage; massive transfusion criterion RBCUnits24h ≥ 10. Analytic n = 559 encounters from 536 unique patients (22 patients with >1 eligible MT encounter; max 3). | p.6, L12–17 |
| 6 (b) | | For matched studies, give matching criteria and number of exposed and unexposed. | Not applicable to the primary analysis (weighting rather than matching). Sensitivity analysis: 1:1 nearest-neighbour matching on the logit of the propensity score with a caliper of 0.2 × SD; matched-pair counts will be reported alongside the matched RD and 95% CI. | p.8, L12–17 (sensitivity 1); p.10, L11–12 |
| 7 | **Variables** | Clearly define all outcomes, exposures, predictors, potential confounders, and effect modifiers. Give diagnostic criteria, if applicable. | Exposure (binary): RBC-only (Plasma Units Total = 0 AND Platelet Units Total = 0) vs Balanced (Plasma Units Total > 0 OR Platelet Units Total > 0). Primary outcome: in-hospital mortality (binary). Secondary: ICU admission (binary), length of stay (continuous, days). Confounders in the PS model (10 covariates): Age at Admission, Sex, First Hemoglobin, Hgb Nadir 24h, First Platelet, First BUN, Variceal Bleed, Had Endoscopy, RBCUnits24h, Admitting Diagnosis subtype (UGIB/LGIB/Unspecified GI, reference = Unspecified GI). Creatinine removed from the analyses due to urine result contamination to the dataset. | p.6, L18–23; p.7, L1–2 |

| Item No. | Topic | STROBE recommendation | Response | Page / Line |
|---|---|---|---|---|
| **8** | **Data sources / measurement** | For each variable of interest, give sources of data and details of methods of assessment (measurement). Describe comparability of assessment methods if there is more than one group. | All variables abstracted from a single institutional source (EHR data warehouse). Creatinine column underwent a data-integrity audit reflecting probable non-serum-sample (most likely spot urine creatinine) contamination in the source extract. | p.6, L9–11; p.7, L10–12 |
| **9** | **Bias** | Describe any efforts to address potential sources of bias. | Confounding by indication addressed by AIPTW with a pre-specified 10-covariate PS and a 6-covariate outcome regression (doubly robust). Selection bias by narrow diagnosis labels addressed by pre-specified broad-diagnosis sensitivity using regex (n=762/727 patients). Unmeasured confounding addressed by E-value for the adjusted RD. | p.7, L13–25; p.8, L1–11 |
| **10** | **Study size** | Explain how the study size was arrived at. | Cohort of all eligible MT encounters in the 5-year window. | p.6, L12–17; p.9, L3–5 |
| **11** | **Quantitative variables** | Explain how quantitative variables were handled in the analyses. If applicable, describe which groupings were chosen and why. | Continuous covariates in the PS model entered as restricted cubic splines. Outcome regression uses the same continuous covariates. Subtype is a 3-level factor (UGIB / LGIB / Unspecified GI, reference = Unspecified GI). Sex coded Male = 1. Hemoglobin reported in g/dL; LOS in days; risk differences in percentage points (pp). | p.7, L21–22 |
| **12 (a)** | **Statistical methods** | Describe all statistical methods, including those used to control for confounding. | Primary: AIPTW estimator targeting the ATE. PS = logistic regression on continuous covariates; IPTW weights w = 1/PS (Balanced), 1/(1−PS) (RBC-only); one-sided 99th-percentile winsorisation on the IPTW weights with pre- and post-winsorisation mean and max reported. Outcome regression: logistic for binary outcomes adjusting for a pre-specified 6-covariate set (First Hemoglobin, Hgb Nadir 24h, Age at Admission, First Platelet, Variceal Bleed, Subtype; gamma GLM with log link for LOS. Inference: non-parametric cluster bootstrap, 2000 resamples, patient-level resampling, percentile 95% CI. NI decided by upper 97.5% CB of RD (RBC-only − Balanced) below the 5-pp margin (3 pp sensitivity). | p.7, L13–25; p.8, L1–11 |

| Item No. | Topic | STROBE recommendation | Response | Page / Line |
|---|---|---|---|---|
| **12 (b)** | | Describe any methods used to examine subgroups and interactions. | No subgroup analyses were planned. | N/A — no subgroup analyses (see p.8, L12–17) |
| **12 (c)** | | Explain how missing data were addressed. | Primary: complete-case on PS covariates with <2% missing (age, sex, Hgb, Hgb Nadir 24h, platelets, BUN, varices, endoscopy, RBC units, subtype). First Creatinine removed due to urine result contamination to the dataset. FirstI NR, Any Anticoagulant, Any Antiplatelet, and First Lactate excluded from the primary PS due to high baseline missingness (≈18%, 46%, 46%, 64% in the MT cohort) and retained for sensitivity analyses. | p.7, L10–12, L18; p.8, L12–17 |
| **12 (d)** | | If applicable, explain how loss to follow-up was addressed. | Not applicable. Primary outcome (in-hospital mortality), ICU admission, and LOS are all in-hospital outcomes ascertained from the same encounter; no post-discharge follow-up is required. | N/A — in-hospital outcomes only (p.7, L1–[illegible] |
| **12 (e)** | | Describe any sensitivity analyses. | (1) 1:1 PS matching, caliper 0.2 × SD logit PS; (2) complete-case adjustment with INR included; (3) broader admitting-diagnosis; (4) first eligible MT encounter per patient only; (5) E-value | p.8, L12–17 |
| **Results** | | | | |
| **13 (a)** | **Participants** | Report numbers of individuals at each stage of study — eg numbers potentially eligible, examined for eligibility, confirmed eligible, included in the study, completing follow-up, and analysed. | Cohort flowchart in Figure 1. Source extract 20,009 inpatient encounters / 13,699 unique patients (2021-01-01 → 2025-12-31) → 3,631 with eligible narrow primary GI bleed admitting diagnosis → 559 encounters meeting RBCUnits24h ≥ 10 from 536 unique patients. | p.9, L3–5; Figure 1, p.19 |
| **13 (b)** | | Give reasons for non-participation at each stage. | Reasons for exclusion documented in the flowchart: age < 18; non-inpatient encounter class; admission outside the 2021–2025 window; Admitting Diagnosis not in the narrow three-label set or broader text-based set covered in sensitivity #3; failed MT criterion (RBCUnits24h < 10). | Figure 1, p.19 |

| Item No. | Topic | STROBE recommendation | Response | Page / Line |
|---|---|---|---|---|
| **13 (c)** | | Consider use of a flow diagram. | Figure 1 CONSORT-style flow diagram. | Figure 1, p.19 |
| **14 (a)** | **Descriptive data** | Give characteristics of study participants (eg demographic, clinical, social) and information on exposures and potential confounders. | Table 1 stratified by exposure. Continuous variables reported as median (IQR), categorical as n (%). | p.9, L5–9; Table 1, p.16–17 |
| **14 (b)** | | Indicate number of participants with missing data for each variable of interest. | Missingness column reported per variable in Table 1. Key missingness in the MT cohort: First Lactate ≈ 64%, Any Anticoagulant / Any Antiplatelet ≈ 46%, First INR ≈ 18%. Variables in the primary PS are all <2% missing. | Table 1 'Missing, n' column, p.16–17 |
| **14 (c)** | | Cohort study — summarise follow-up time (eg, average and total amount). | Length of stay is reported by exposure arm as a descriptive summary; median and IQR appear in Table 1. | Table 1 (LOS), p.17 |
| **15** | **Outcome data** | Cohort study — report numbers of outcome events or summary measures over time. | Unadjusted event counts: in-hospital mortality 6/505 (1.2%) RBC-only vs 8/54 (14.8%) Balanced (narrow cohort). Broad-cohort counterpart: 20 deaths in n=749 analytic. ICU admission and LOS reported per arm in Table 2. | p.9, L7–9; Table 1, p.17 |
| **16 (a)** | **Main results** | Give unadjusted estimates and, if applicable, confounder-adjusted estimates and their precision (eg, 95% CI). Make clear which confounders were adjusted for and why they were included. | Unadjusted RD (mortality, RBC-only − Balanced) = −13.6 pp. Adjusted AIPTW point estimate, 95% percentile cluster-bootstrap CI, and upper 97.5% confidence bound to be read from results/ primary_estimates.csv after the 2026-04-26 rerun. Confounder rationale: the 10-covariate PS captures the strongest clinical drivers of plasma/platelet activation (illness severity, anaemia depth, coagulation surrogate via platelets, AKI surrogate via BUN, anatomy via subtype and varices, intensity of bleeding via RBCUnits24h). 6-covariate outcome-regression set chosen pre-outcome-modelling on the basis of post-weighting SMD ≥ 0.2 (First Platelet, Variceal Bleed, Subtype) plus established mortality predictors (First Hemoglobin, Hgb Nadir 24h, Age at Admission). | p.9, L21–25; p.10, L1–3; Table 2, p.18 |

| Item No. | Topic | STROBE recommendation | Response | Page / Line |
|---|---|---|---|---|
| **16 (b)** | | Report category boundaries when continuous variables were categorized. | Not applicable — no continuous variable is categorised in the primary analysis. Subtype, the only categorical confounder, is a fixed 3-level factor (UGIB / LGIB / Unspecified GI, reference = Unspecified GI). | N/A — no continuous variable categoris[illegible] |
| **16 (c)** | | If relevant, consider translating estimates of relative risk into absolute risk for a meaningful time period. | All primary effect estimates are reported on the absolute risk-difference scale (percentage points), consistent with the pre-specified NI margin. Relative measures (RR, OR) are not used for the primary inference. | Reported as absolute RD throughout (p.9, L23; Table 2, p.18) |
| **17** | **Other analyses** | Report other analyses done — eg analyses of subgroups and interactions, and sensitivity analyses. | Broad-cohort sensitivity currently shows RD = −0.1717 with NI demonstrated at both 5-pp and 3-pp. | p.10, L9–18; Figure 4, p.22; Table 2, p.18 |
| **Discussion** | | | | |
| **18** | **Key results** | Summarise key results with reference to study objectives. | Discussion opens with the adjusted RD and upper 97.5% CB relative to the pre-specified 5-pp and 3-pp margins, and the consistency of the conclusion across sensitivity analyses. NI claim stated with both point estimate and upper bound; no superiority claim from an NI test. | p.10, L19–25; p.11, L1–3 |

| Item No. | Topic | STROBE recommendation | Response | Page / Line |
|---|---|---|---|---|
| **19** | **Limitations** | Discuss limitations of the study, taking into account sources of potential bias or imprecision. Discuss both direction and magnitude of any potential bias. | Six limitations are acknowledged. First, plasma and platelet units are recorded as whole-encounter totals rather than 24-hour counts, precluding computation of acute RBC:plasma:platelet ratios; however, 67% of patients received their entire RBC load within 24 hours, so encounter totals approximate acute resuscitation. Second, the 9:1 exposure imbalance (505 RBC-only vs 54 balanced) limits precision: the effective balanced-arm sample size is approximately 30 (Kish ESS), and six of ten covariates retained residual imbalance (\|SMD\| 0.14–0.29) after weighting despite the doubly robust AIPTW estimator. Third, admission creatinine was excluded from the propensity model due to non-serum sample contamination in the source extract; first platelet count, which differed substantially between arms, is included in the outcome regression to partially compensate. Fourth, as a single-centre retrospective study, unmeasured confounding cannot be excluded, though the E-value of 4.80 for the non-inferiority bound indicates the required confounder strength is implausibly high. Fifth, the temporal distribution of RBC delivery within 24 hours was not captured, limiting inference about rate-dependent dilutional coagulopathy. Finally, findings reflect a single institution and may not generalise to settings with different massive-transfusion triggers, case mix, or endoscopic [illegible] | p.13, L1–27; p.14, L1–13 |
| **20** | **Interpretation** | Give a cautious overall interpretation of results considering objectives, limitations, multiplicity of analyses, results from similar studies, and other relevant evidence. | No randomized trial has directly compared RBC-only versus balanced component therapy in adults meeting massive transfusion criteria for GI hemorrhage. Using doubly robust propensity weighting across 559 encounters, we demonstrate non-inferiority of RBC-only at both pre-specified 5-pp and 3-pp mortality margins across five sensitivity analyses, providing the first systematic evidence to support decoupling trauma resuscitation ratios from GI hemorrhage management. | p.11, L4–28; p.12, L1–27; p.14, L14–20 |

| Item No. | Topic | STROBE recommendation | Response | Page / Line |
|---|---|---|---|---|
| **21** | **Generalisability** | Discuss the generalisability (external validity) of the study results. | Single tertiary-care center; admissions 2021–2025; transfusion practice and access-to-endoscopy patterns may differ at community centres; cohort dominated by Unspecified-GI subtype (446 of 559), so generalisation to populations with a higher UGIB share (e.g., variceal-bleed-heavy centres) requires the subtype-interaction reanalysis flagged under Item 12(b). | p.14, L11–13 |
| **Other information** | | | | |
| **22** | **Funding** | Give the source of funding and the role of the funders for the present study and, if applicable, for the original study on which the present article is based. | This research received no specific grant from any funding agency in the public, commercial, or non-profit sectors. Institutional computational infrastructure was provided through the umbrella data-platform protocol (CMTT-PROJ 210925). | p.1, L14; p.15, L6–8 |
