## Supplementary material for "Non-inferiority of a red-blood-cell–only transfusion strategy compared with balanced resuscitation in adults with massive gastrointestinal haemorrhage: a propensity-score–weighted cohort study": CONSORT-NI extension

### CONSORT-NI adapted reporting table

| Item No. | Topic | CONSORT-NI extension requirement | Observational adaptation / Our response | Page / Line |
|---|---|---|---|---|
| **Title and abstract — NI-specific additions** | | | | |
| **1a** | **Title** | CONSORT-NI extension: Identify the study as non-inferiority (or equivalence) in the title. | Title explicitly identifies the study as a non-inferiority cohort analysis (e.g., "Non-inferiority of RBC-only versus balanced transfusion in adults with massive gastrointestinal hemorrhage: a retrospective cohort study"). The phrase "non-inferiority" is required by CONSORT-NI item 1a even though the design is observational. | p.1, L1–2; p.2, L6–8 |
| **1b** | **Abstract** | CONSORT-NI extension: The abstract should state the non-inferiority hypothesis, the pre-specified margin, the analysis population(s), and the conclusion in terms of the confidence-interval bound relative to the margin. | Structured abstract carries the NI hypothesis (RBC-only vs Balanced), the pre-specified 5-pp absolute-RD primary margin (one-sided α = 0.025) and 3-pp sensitivity margin. | p.2, L1–25 (NI hypothesis L6–8; margins L12–13; conclusion L17–18) |
| **Introduction — NI-specific additions** | | | | |
| **2a** | **Background / rationale** | CONSORT-NI extension: Provide the rationale for adopting a non-inferiority (rather than superiority) design. | NI rationale stated explicitly in Introduction: balanced 1:1:1 resuscitation in trauma addresses the trauma triad, which is rarely present in MT-requiring GI hemorrhage. The clinically relevant question is therefore whether plasma/platelets can be safely withheld in this population a non-inferiority framing, not a superiority framing. | p.5, L15–18 |
| **2b** | **Specific objectives / hypotheses** | CONSORT-NI extension: State the non-inferiority hypothesis with the pre-specified margin, the direction of the comparison, and the side of the test (one-sided vs two-sided). | Pre-specified NI hypothesis: RBC-only is non-inferior to Balanced with respect to in-hospital mortality if the upper bound of the one-sided 97.5% confidence interval for the absolute risk difference (RBC-only − Balanced) lies below 5 percentage points. Direction of inferiority: RBC-only worse. Test: one-sided, α = 0.025. Pre-specified sensitivity margin: 3 pp. | p.5, L15–21; p.7, L3–9 |

| Item No. | Topic | CONSORT-NI extension requirement | Observational adaptation / Our response | Page / Line |
|---|---|---|---|---|
| **Methods — NI-specific additions** | | | | |
| **5** | **Interventions / exposures** | CONSORT-NI extension: Describe the comparator and its assumed effect carefully — non-inferiority assumes the comparator preserves a known benefit ("assay sensitivity"). State the assumed benefit and how it was derived. | Comparator (Balanced strategy) is the de facto trauma-extrapolated standard in this study. The comparator-effect is the historical MT-in-GI-hemorrhage mortality range (1–15% in this cohort; PROPPR 24-h mortality differences of ~10 pp in trauma; TRIGGER 4-pp RDs in UGIB) used to justify the 5-pp margin. Assay-sensitivity surrogate: the broad-cohort sensitivity and the unadjusted vs adjusted comparison together demonstrate that the estimator responds in the expected direction to known confounding patterns (illness severity). | p.6, L18–23; p.7, L3–9 |
| **6a** | **Outcomes — pre-specified** | CONSORT-NI extension: Pre-specify the primary outcome on which non-inferiority will be decided. Multiple primary outcomes are discouraged in NI trials. | Single pre-specified primary outcome: in-hospital mortality (binary). Rationale: complete in all 559 cohort encounters; clinically paramount; directly comparable to PROPPR / TRICC / TRISS / TRIGGER. Secondary outcomes (ICU admission, length of stay) are not NI-tested; reported with adjusted RD and 95% CI (binary) or weighted median difference with bootstrap CI plus gamma-GLM sensitivity (LOS). | p.7, L1–2; p.7, L3–9 |
| **7a** | **Sample size / margin** | CONSORT-NI extension: Justify the chosen non-inferiority margin clinically and statistically. State the assumed event rates, power, and α used in the sample-size calculation. | Margin = 5 pp absolute RD (RBC-only − Balanced), one-sided α = 0.025; 3-pp sensitivity margin. Justification: (i) baseline in-hospital mortality in this cohort is 1–15%, so a 5-pp absolute increase is clinically meaningful and would require offsetting benefit to accept; (ii) 5 pp is conservative relative to the unadjusted crude RD of −13.6 pp; (iii) previous literature (PROPPR) powered for ~10-pp differences (Holcomb JAMA 2015); TRICC 5.5-pp NI margin (Hébert NEJM 1999); TRISS 6-pp (Holst NEJM 2014); TRIGGER and Villanueva considered 4-pp RDs clinically important; 3–5 pp is the conventional range for absolute-RD NI margins on mortality in transfusion / critical-care trials. | p.7, L3–9; p.13, L11–18 |

| Item No. | Topic | CONSORT-NI extension requirement | Observational adaptation / Our response | Page / Line |
|---|---|---|---|---|
| **11** | **Blinding** | CONSORT-NI extension: Where outcome assessors cannot be blinded (typical for an observational design), describe steps to limit differential ascertainment, which threatens NI conclusions because biased ascertainment can falsely shrink the gap between arms. | Outcome (in-hospital mortality) is ascertained from a single institutional source independent of transfusion-strategy classification. Differential ascertainment is therefore implausible for the primary outcome. ICU admission and LOS are also recorded by the same source. Exposure ascertainment is mechanically defined from transfusion totals (Plasma Units Total, Platelet Units Total); no blinding involved because exposure status is a property of the EHR record, not of any human classifier. | p.13, L19–23 |
| **12a** | **Statistical methods — NI** | CONSORT-NI extension: Specify (i) the statistical method, (ii) the analysis populations (ITT and per-protocol), and (iii) the rule for declaring non-inferiority (which CI bound, vs which margin). | Primary estimator: AIPTW (doubly robust) for the ATE; PS via logistic regression with restricted cubic splines (3 knots at 10th/50th/90th percentiles, Harrell); IPTW weights one-sided 99th-percentile winsorised; outcome regression with a pre-specified 6-covariate set. Inference: non-parametric cluster bootstrap, 2000 resamples, patient-level (PatientKey). Non-inferiority declared when the upper 97.5% percentile-bootstrap bound for the absolute RD (RBC-only − Balanced) lies below the 5-pp primary margin (3-pp for the sensitivity margin). If the bound exceeds the margin, the result is "inconclusive", not "inferior"; if upper bound falls below 0 then "consistent with potential superiority". | p.7, L13–25; p.8, L1–11 (NI decision rule p.8, L6–8) |
| **12a-ITT** | **ITT / per-protocol — observational analogue** | CONSORT-NI extension: Both ITT and per-protocol analyses should be reported in NI trials; the NI conclusion should be consistent across them. | In an observational cohort, the ITT vs per-protocol distinction translates to (a) primary AIPTW on the full eligible cohort using the as-observed exposure (analogue of ITT) and (b) pre-specified sensitivity analyses that restrict the analytic sample or sharpen the comparison (analogue of per-protocol). (1) PS matching; (2) complete-case with INR; (3) broad-cohort; (4) first MT encounter per patient; (5) E-value (§4.4 #5). NI conclusion is reported as robust only if it holds across all rows; details discussed in Results and Discussion. | p.6, L15–17 (ITT-analogue); p.8, L12–17 (per-protocol analogue) |
| **12b** | **Additional analyses** | CONSORT-NI extension: Pre-specify subgroup and interaction analyses; flag any post-hoc analyses. | No subgroup analyses performed, pre- or post. | p.8, L12–17 |
| **Results — NI-specific additions** | | | | |

| Item No. | Topic | CONSORT-NI extension requirement | Observational adaptation / Our response | Page / Line |
|---|---|---|---|---|
| **13a** | **Participant flow** | CONSORT-NI extension: Report the analysis populations separately (ITT and per-protocol equivalents). For observational designs, report the full cohort and any restricted-cohort sensitivity counts. | Figure 1 reports the cohort flow from the source extract (20,009 encounters / 13,699 patients) to the analytic primary cohort (n=559 / 536 patients) and to the analytic broad-cohort sensitivity (n=749 after complete-case from 762 eligible). | p.9, L3–5; Figure 1, p.19; broad cohort p.10, L13–16 |
| **17a** | **Outcomes and estimation** | CONSORT-NI extension: Report a forest-style figure or table showing the point estimate and confidence interval for the primary outcome, alongside the non-inferiority margin. The CI's position relative to the margin should be readable at a glance. | Figure 2 showing the AIPTW adjusted RD for in-hospital mortality with 95% percentile-bootstrap CI, the upper 97.5% confidence bound, and vertical reference lines at 0 pp (null), 3 pp (sensitivity margin), and 5 pp (primary margin). | p.9, L21–25; p.10, L1–3; Figure 4, p.22; Table 2, p.18 |
| **17b** | **Binary outcomes** | CONSORT-NI extension: For binary outcomes, present absolute and relative effect sizes with confidence intervals. | Adjusted RR or OR with 95% CIs reported as secondary metrics in Table, the NI decision is made on the RD scale because the pre-specified margin is on that scale. | Table 2, p.18; p.9, L21–25; p.10, L1–3 |
| **18** | **Ancillary / sensitivity analyses** | CONSORT-NI extension: Report each pre-specified sensitivity analysis with the same effect-and-CI format as the primary, so the reader can judge robustness of the NI conclusion. | NI demonstrated at both 5-pp and 3-pp margins in both R and Python implementations. The full sensitivity table is included in the Results section and the matching forest figure. | p.10, L9–18; Figure 4, p.22; Table 2, p.18 |
| **Discussion — NI-specific additions** | | | | |
| **20** | **Limitations — NI-specific** | CONSORT-NI extension: Discuss factors that bias an NI estimate toward the null (i.e., toward a false NI conclusion). These include misclassification of exposure or outcome, drift toward usual care, and any analytic feature that shrinks observed differences. | (1) Component-data asymmetry: Plasma Units Total and Platelet Units Total are whole-encounter totals, not 24-h totals, so the Balanced arm includes patients whose plasma/platelet receipt was not MT-related. (2) Residual confounding from variables excluded from the primary PS (lactate, INR, anticoagulant/antiplatelet); bounded by the pre-specified E-value. (3) Outcome ascertainment limited to in-hospital, does not capture out-of-hospital or 30-day mortality. | p.13, L1–27; p.14, L1–13 |

| Item No. | Topic | CONSORT-NI extension requirement | Observational adaptation / Our response | Page / Line |
|---|---|---|---|---|
| 21 | **Generalisability — NI-specific** | CONSORT-NI extension: Discuss whether the trial population resembles the historical control population on which the comparator's assumed benefit rests (a threat to assay sensitivity). | Cohort is dominated by Unspecified-GI subtype (446/559 narrow primary cohort); UGIB (64) and LGIB (49) shares are modest. Variceal bleeders are present (Variceal Bleed indicator) but not numerically dominant. Trauma-derived 1:1:1 comparator effect (PROPPR) was estimated in injury populations with universal coagulopathy at presentation, a population materially different from this cohort, where coagulopathy is the exception. This mismatch is the assay-sensitivity threat for the NI design and is discussed explicitly in Discussion. | p.14, L11–13 |
| 22 | **Interpretation — NI-specific** | CONSORT-NI extension: State the conclusion explicitly in terms of the confidence-interval bound and the pre-specified margin. Distinguish "non-inferior" from "inconclusive". Do not claim superiority from an NI test alone. | NI conclusion stated with (i) the adjusted point estimate, (ii) the upper 97.5% confidence bound, (iii) the pre-specified 5-pp primary margin (and 3-pp sensitivity margin); if the upper bound is below the margin, the conclusion is "non-inferior"; if it crosses the margin, the conclusion is "inconclusive", not "inferior". Superiority is not claimed from this NI design unless the upper bound falls below 0, and even then framed as "consistent with potential superiority", not "superior". | p.10, L1–3; p.14, L14–20 |
| **Other information** | | | | |
| 23 | **Registration / pre-specification** | CONSORT-NI extension: NI trials should be pre-registered with the NI margin specified at registration. Where observational studies cannot be registered in trial registries, the analogue is dated pre-specification of the analysis plan with the margin and primary estimand. | Analysis plan pre-specified 2026-04-18 (primary outcome, NI margins, primary AIPTW, sensitivity-analysis list). Subsequent amendments documented and dated.Pre-specification record committed alongside code; can be supplied to editors on request. | p.1, L16–18; p.6, L4–7; p.15, L9–13 |
| 24 | **Protocol / analysis plan availability** | CONSORT-NI extension: The protocol with the NI design rationale and margin justification should be available to reviewers and readers. | Both can be made available on request and will be referenced in the Data Availability statement. | p.15, L14–19 |
